# It’s the very time to learn a pandemic lesson: why have predictive techniques been ineffective when describing long-term events?

**DOI:** 10.1101/2020.06.01.20118869

**Authors:** D. Kovriguine, S. Nikitenkova

## Abstract

We have detected a regular component of the monitoring error of officially registered total cases of the spread of the current pandemic. This regular error component explains the reason for the failure of a priori mathematical modelling of probable epidemic events in different countries of the world. Processing statistical data of countries that have reached an epidemic peak has shown that this regular monitoring obeys a simple analytical regularity which allows us to answer the question: is this or that country that has already passed the threshold of the epidemic close to its peak or is still far from it?

## Summary

The current epidemic has spread almost unhindered throughout the world, becoming a pandemic. Scientists very quickly identified a new type of virus and sequenced the virus genome [1], which will undoubtedly reduce the time before the appearance of an effective vaccine.

Unfortunately, medicine had neglected to learn the lessons of the last major epidemic in 1968. Many people had not got elementary knowledge on sanitary counteraction to the epidemic even in our twenty-first century [2]. Sanitary services in many countries have belated with preventive actions. We should exclude these faults in the future.

Today, in the era of “fantastically” developed communications and computational methods, mathematicians should contribute to the combat against the pandemic. First of all, people desire to know about future epidemic events in terms of where, how much and when. This information creates assurance in society, some adequate understanding and effective reaction on current events. Studying the history of the pandemic in detail will undoubtedly contribute to fruitful strategic thinking.

The practice of recent months has shown that well-tested and well-known mathematical methods for processing statistical data directly used to describe future events of the spread of a pandemic have been ineffective in solving such a flagrantly important and urgent problem. Meanwhile, it is no secret that the aggregated dynamics of a pandemic is very simple, it is described quite fully by the solution of a well-known logistic equation, not so important whether it is discrete or differential. Neither the more developed versions of this mathematical model, based on the introduction of new unknown variables nor the tricks of the multiparameter modification of the logistic equation help the case. In any case, the simulation gives a satisfactory result only on a small-time horizon, from a force limited to one week. If so, then we are dealing not with predictive, but with monitoring models, which, for all their value, are still useless for developing medium-term and strategic action plans to overcome the pandemic. Readers can find examples of enhanced monitoring in the sites [3], [4], etc. Other promising predictive technologies based on graph theory, percolation theory, and stochastic processes turned out to be too clumsy or incapable to produce a clear and concrete result on an urgent issue. In other words, the available methods for processing statistical data were found to be helpless in solving an important specific problem. The paradox is the apparent inefficiency of using the known methods of processing statistical data to obtain an adequate quantitative result, even though, it would seem, everything about the dynamics of the pandemic is known qualitatively in advance.

## Method

Let’s try to understand the main reasons for such an unfortunate failure and try to formulate the principles for overcoming the current difficult situation.

First of all, what does an epidemic look like? In the dynamics of development and spread, the epidemic is similar to a forest fire. With fire, everything is understood a little easier due to the obviousness of this phenomenon. No one is trying to delve into the physics of combustion and the nature of fire, for people the expectations of ignition of adjacent sections of the forest, threats to human settlements are obvious, the assessment of the forces and means necessary to extinguish a fire is almost foreseeable. The epidemic does not have such striking external manifestations, so it is insidious and dangerous especially for ignorant people, in which, as a rule, there is no shortage. Besides, decisive actions taken, say, to extinguish a fire, are completely inapplicable if you try to try them on to combat the epidemic. In the absence of vaccination and other effective medical countermeasures to the spread and development of the epidemic, quarantine seems to be the most effective means, like fifty years ago and earlier. Self-isolation and social distancing are effective only when each member of the society realizes that any action, even if it is legitimate, borders on risk.

To understand the reason for the failures of mathematical modelling, we first turn our attention to the statistical data used to make the forecast, since this forecast directly depends on them. It is not a revelation that, for completely natural reasons, statistics are not necessarily flawless. Note that, in contrast to the experimental data that are dealt with, say, in physical experiments, data on the epidemic situation cannot be redundant, but only insufficient. Indeed, it is difficult to imagine a situation where redundant data regularly appears in the monitoring summary of the number of newly detected cases, since the case is too delicate to allow such sloppiness. Most likely, we should expect that the data will be underestimated due to a lack of information. Therefore, it is natural to assume that the data contains a regular error component with a deficiency. Indeed, comprehensive pandemic data can only be obtained a posteriori, but life requires reliable a priori information. To achieve this goal, it is necessary to identify, evaluate and study the mentioned regular component of the error, using the statistics of those countries that have already reached a peak – the stationary level of the epidemic dynamics.

Let us take a logistic pattern for the predicted result since it requires a minimum of information for modelling, i.e., the initial number of cases, the final peak number, and the number of days from the outbreak to its peak. One can use appropriate data to build these logistic curves, for example from the site [5]. Rearrange time series from data tables as an autocorrelation function *N*_*i*+1_ = *f_c_*(*N_i_*), where *N_i_* is the number of total cases per day number *i* in the country *c*. Following the least-squares method, we determine polynomial approximations *F_c_* to the tabular functions *f_c_*. Numerical experiments confirm the sufficiency of the quadratic approximation of functions *f_s_* from the argument *N_i_*. So, the dynamic model takes the form of an analytical point mapping 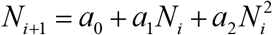. Here, *a_i_* are the parameters determined from the tables. The transition time from the state *N_i_* to the state *N*_*i*+1_ is a day, but it does not play a significant role at large numbers. After using simple selection criteria, the obtained data appears as tables and graphs [6].

Suppose that the temporary evolution of the epidemic in countries possessing high-quality statistics is adequately described by the solutions of the logistic equation 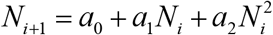 when the phase point of this dynamical system passes from the unstable stationary state *N* = 0 into the steady-state *N* = *N*\*, where *N*\* Is the positive root of the quadratic equation *N*\* = *a*_0_ + *a*_1_*N*\* + *a*_2_ *N*^*2^. The steady-state *N*\* corresponds to almost zero increase in disease or the epidemic peak. At the time of achieving this stable equilibrium, the logistic equation is no longer sufficient to describe the evolution of the disease in a developed human society. The fate of infected people depends on the quality of quarantine measures, medical care, etc. There is no doubt that, theoretically, the epidemic can develop for the second and third time, etc., in an unpredictable way, if neglecting appropriate anti-epidemic cares.

Let the infection rate be a specific value for each territory or country. It is easy to guess that the parameter *a*_0_ should tend to zero due to elementary statistical properties. Thus, we can interpret the relative indicator *a*_0_/*N*^*^ as the quality of statistical data. Always a positive parameter *a*_1_ determines the maximum spread of the epidemic. Let it characterize the measure of social hygiene inherent in a particular country. The critical parameter determining a stable steady-state *N*^*^is this *a*_2_. Note that abstract theories connect this parameter with the concept of intraspecific competition, but in our case, an attempt to interpret this indicator will lead to nothing. It remains to be assumed that this negative parameter is an indicator of medical and other useful actions to counteract, spread, and develop the epidemic.

Now let’s look at a specific example of the epidemic history in one of the countries that have reached the peak of the epidemic. Let it be the most affected European country today, Spain, shown in Fig. 1.

**Fig. 1.**
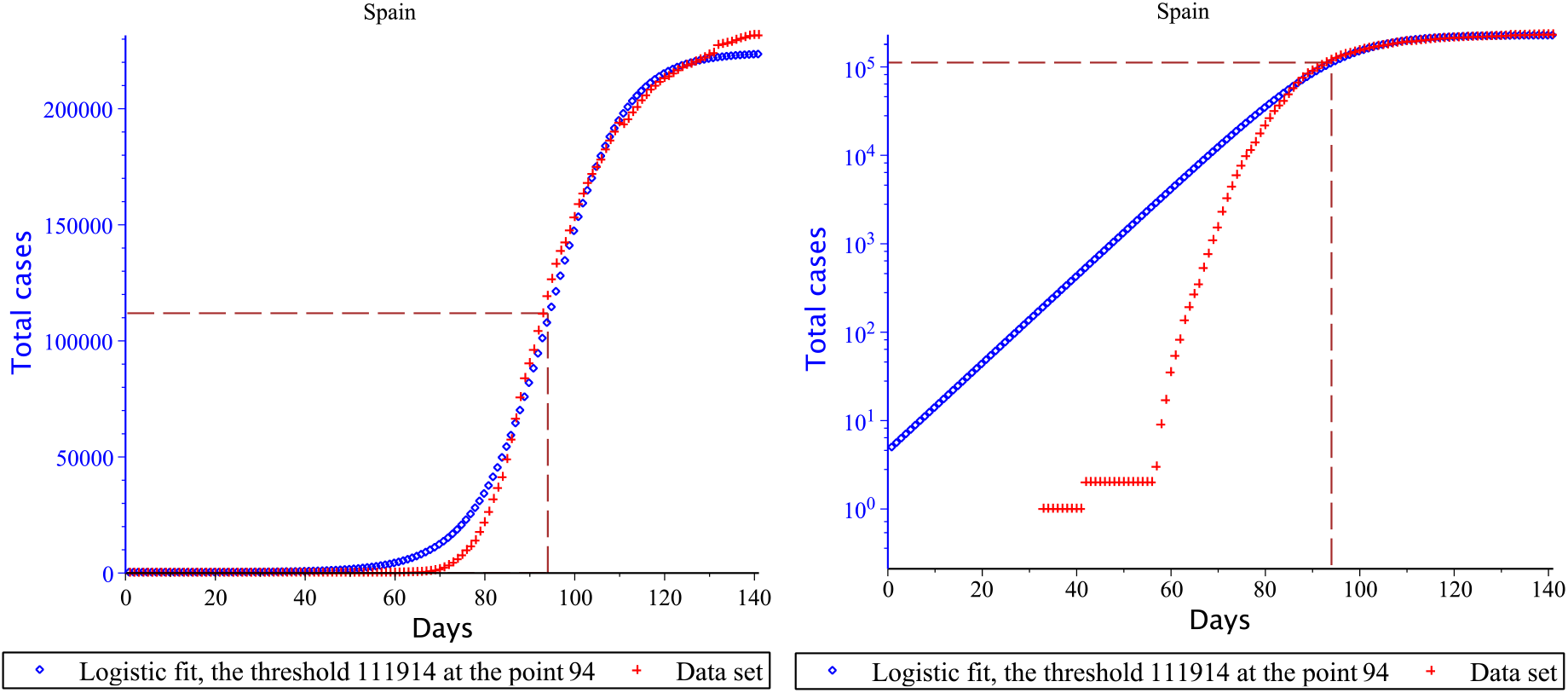
The history of the epidemic in Spain. The red symbols indicate the total number of officially registered cases of infection by day, starting from the beginning of the epidemic in this country. The blue symbols represent the forecast line. The right graph is the same history presented in logarithmic scale.

The above graphs show that the discrepancy between real data and the forecast is most significant, starting from the early stages of the epidemic until the inflexion point of the logistic curve, which is appropriate to call a threshold. In the figures, this threshold is indicated by a dotted line. Now let’s try to evaluate the absolute and relative forecasting error graphically, as shown in Fig. 2.

**Fig. 2.**
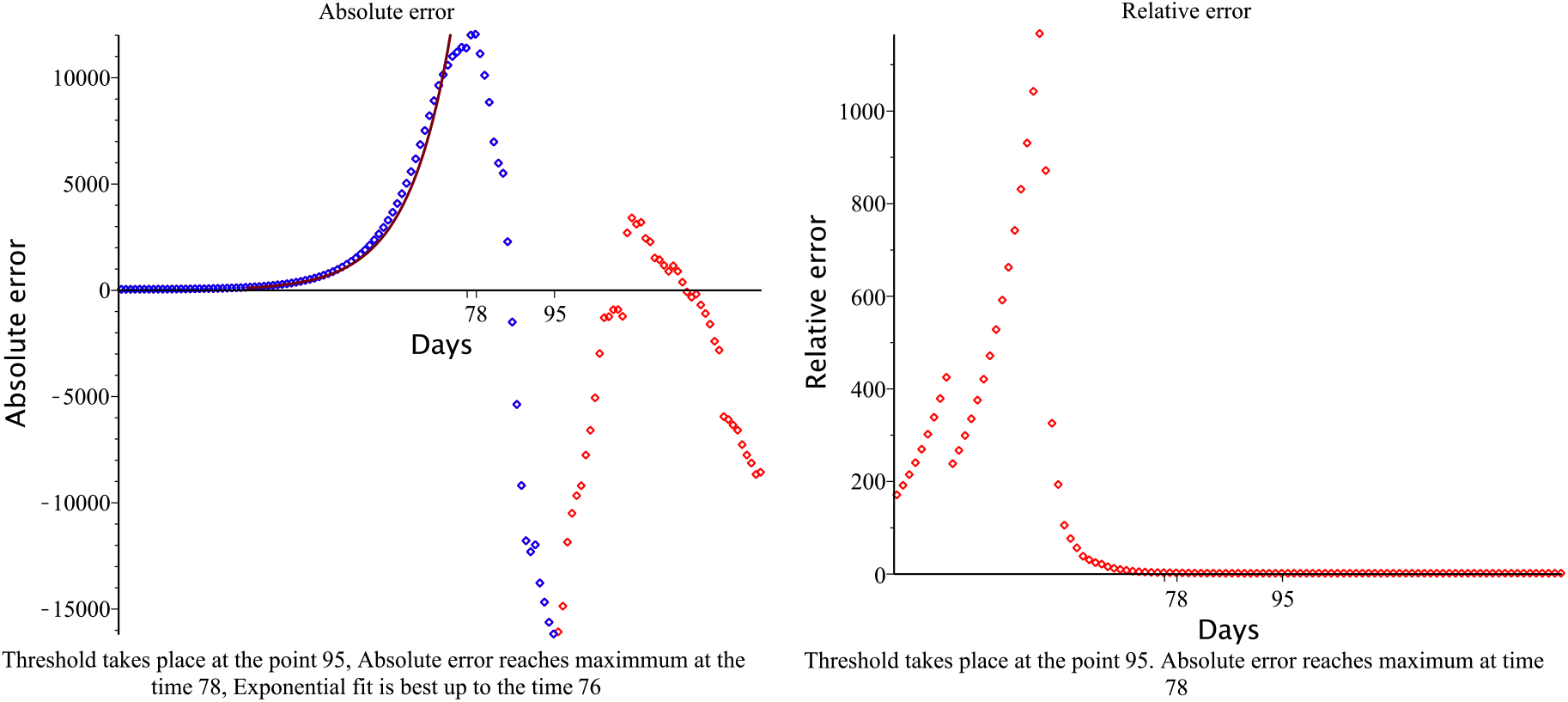
Absolute and relative forecast errors.

This illustration shows that absolute error is colossal throughout the history of the epidemic. The relative error becomes acceptable when the absolute error has passed its maximum value. Fortunately, the actual data, starting from the beginning of the epidemic and almost to the point of the specified maximum, is well described by the exponential dependence shown in the left part of Fig. 2. This fact is not a revelation: the behaviour of real data over time shown in graphs is not a specific feature of the history of the epidemic in Spain alone. The authors of some works on the problem of the pandemic have noticed this pattern, but link this regular exponential component of the error with useful medical intervention in the course of the early stages of the epidemic [7]. However, it is easier to assume that this unavoidable regular error is due only to the specifics of data monitoring. Moreover, the analytical approximation of the input data set continues further, starting from the point of maximum absolute error up to the inflexion point [8], [9], [10]. Figure 3 shows the power fit on the indicated time interval.

**Fig. 3.**
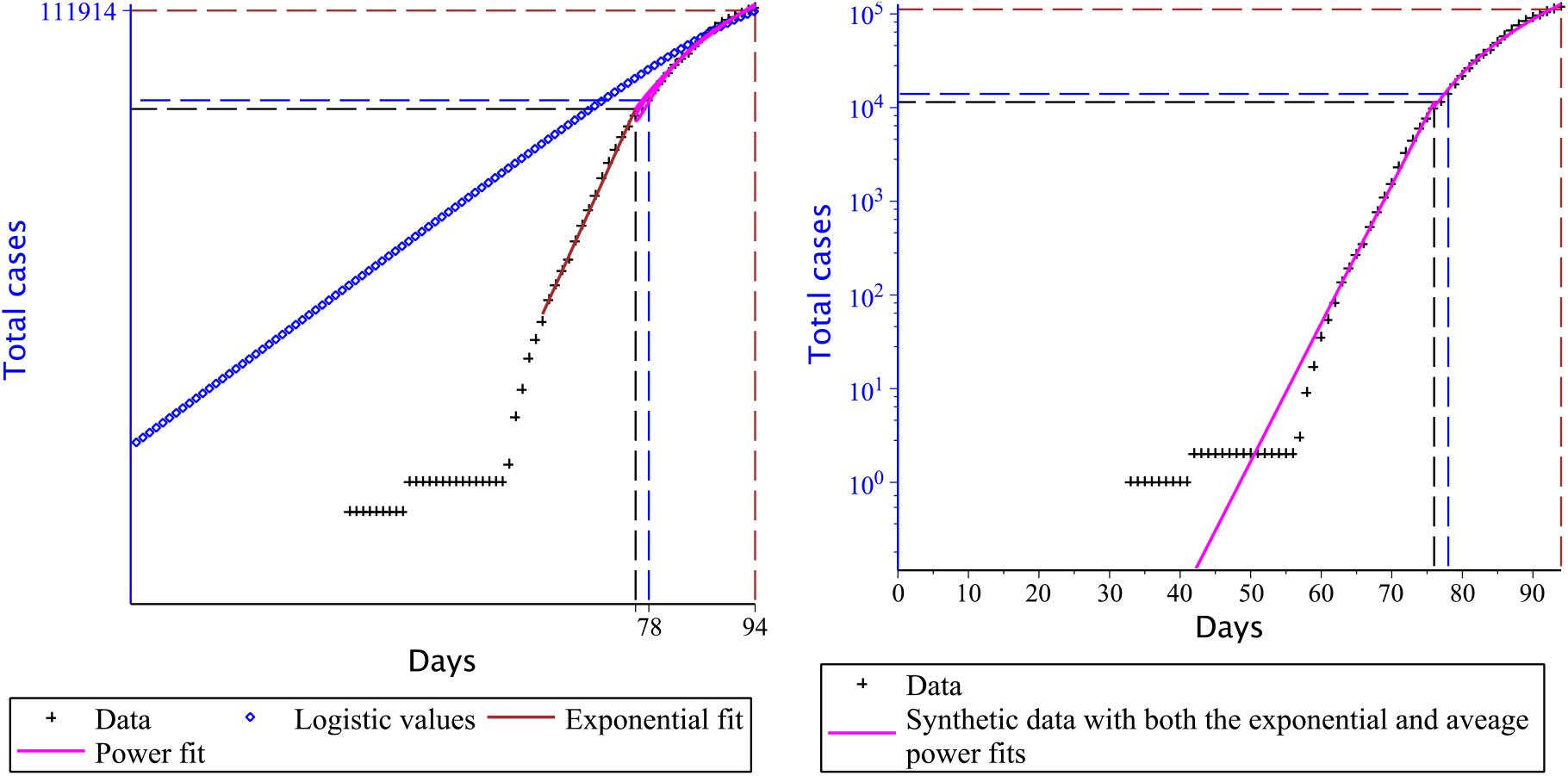
Exponential and power approximation of input data. The graph on the right shows the possibility of replacing the monitoring data with approximating functions.

These graphs show that real input data can be replaced by approximating functions without any particular error. Note that beyond the inflexion point, and up to the peak of the epidemic, the predicted and real data, as a rule, practically coincide. The explanation is simple: when crossing the inflexion point, the growth rate of registered infections decreases, while the maximum effort is involved in solving the monitoring problem. If we repeat the calculation procedure using any suitable technique for processing statistical data using the indicated exponential and power-fit approximating functions, the result will be close to what is achieved using real input data.

## Results

We can repeat the above arguments and calculations using other data on the history of the epidemic in other countries, for example, Germany, Italy, Turkey, France, and many others. The pattern in the input data in question will be the same. This means that epidemic event monitoring services in many countries operate similarly, which causes the inevitable regular error component to occur. Of course, there are exceptions: there are data for some countries for which the analytical analysis described in this paper cannot be used. Such countries, among the most affected by the pandemic, primarily include China, whose monitoring services provided data that could not be processed in the above aspect. Also, in this aspect, countries such as Ecuador, Japan, Slovakia, Guinea-Bissau, Kosovo are not amenable to statistical processing. In some other countries, as a rule, with a few registered cases, the causal principle is formally violated. This means that monitoring data overtakes in time the data calculated by the logistic forecast curve. Since in reality this principle is never violated, it remains to be assumed that the monitoring data provided by such countries is incomplete. If desired and zealous, this error is not difficult to correct, assuming that the epidemic could have begun somewhat earlier than the officially declared date. There are some countries, also, as a rule, with some recorded cases, where the forecast data surprisingly practically coincide with the monitoring data. These countries include Ireland, Iceland, New Zealand.

We assume that the regular component of the monitoring error has a place to be. Could this information have a positive effect on the reliability of predicting future epidemic events in other countries that have not yet reached the peak of the epidemic? If we take into account the evolution of the indicated regular error component in a dynamic way, that is, ad-just the input data appropriately, this problem will not completely disappear. The parameters of the exponential and power functions that approximate the input data are unknown a priori.

The history of countries that have reached the peak of the epidemic is of fundamental value. This value is manifested in the appearance of a pattern in the so-called virtual time delay of monitoring data. The indicated delay time is determined as follows. Mentally, we draw a horizontal line on the graph, which is shown on the left in Fig. 3. We determine the intersection points of this line with the forecast logistics curve and graphs of functions that approximate the input data. The time difference at the indicated intersection points determines the virtual delay time of the monitoring data. In other words, if you move the monitoring points from right to left by the desired time interval, then all the points will be on the logistic curve, which will lead to a correct prediction, even if you do not use all the new data obtained, but only a small part of them. In Fig. 4 presents virtual time delays for a representative number of countries that have reached the peak of the epidemic. These are countries such as Spain, Italy, Germany, Turkey, France, Belgium, Netherlands, Switzerland, Portugal, Israel, Austria, Denmark, Serbia, Czech Republic, Norway, Finland, Luxembourg, Hungary, Croatia, Macedonia, Cuba, Estonia, Tunisia, Latvia. Monitoring data from these countries we have selected according to the following criteria:

1. there are both the exponential and power approximating fits;
2. the virtual time delay is defined;
3. replacing the real input data with approximating functions creates an error of not more than 33 per cent;
4. a country has fetched its epidemic peak to the current date for at least a week ago.

Figure 4 demonstrates an obvious fact that the higher the epidemic threshold, then the greater the virtual time delay.

**Fig. 4.**
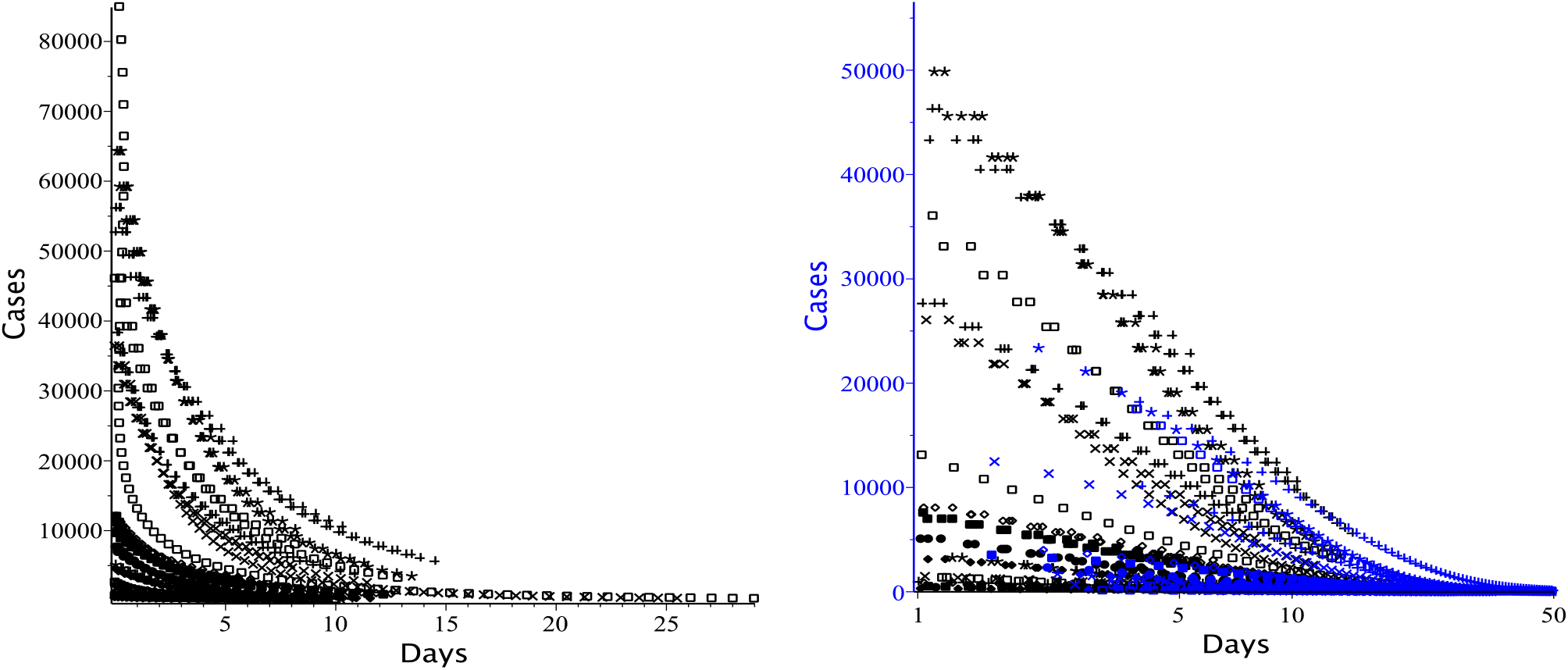
Virtual time delay of monitoring data. The graph on the left shows the main part of the time delay calculated using a power-law approximation of the input data. In the illustration to the right of the main part, an insignificant part of the time delay, which is calculated using the exponential approximation of the monitoring data, is added.

Now consider the nature of the virtual time delay in key countries that have crossed the threshold of the epidemic, but have not yet reached its peak. Figure 5 shows that monitoring data from Canada, the United Kingdom, and Saudi Arabia generally obey the patterns of virtual delay. The time delays of Peru, and especially Russia, are uncharacteristic of the event approaching the peak of the epidemic.

**Fig. 5.**
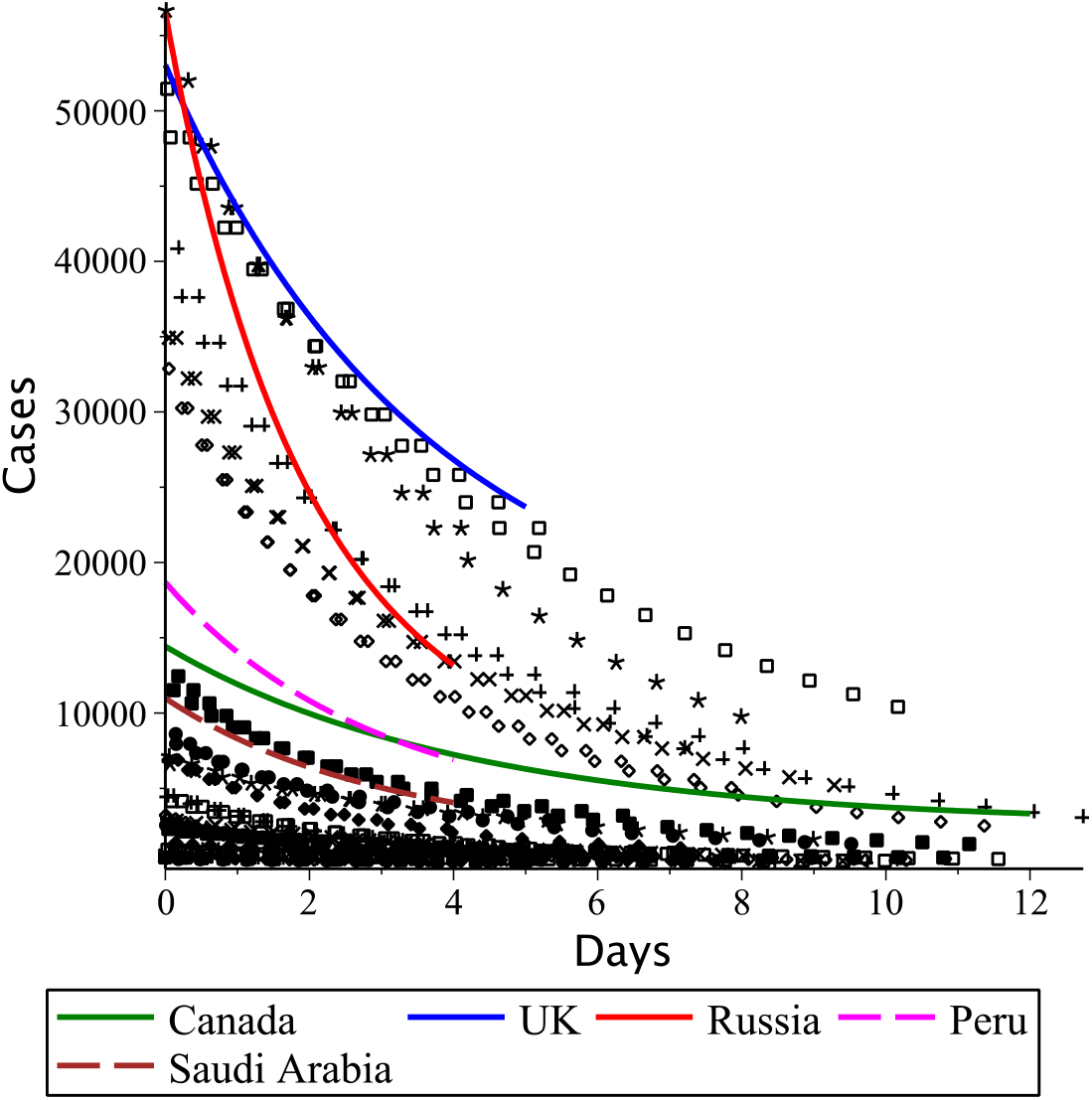
Virtual time delay of epidemic monitoring data in Canada, the United Kingdom^†^, Peru, Saudi Arabia and Russia, which have crossed the epidemic threshold but have not yet reached its peak. We have used the same illustration is as in Fig. 4 on the left as a background for visual comparison.

Unfortunately, the above analysis cannot be applied to US monitoring data, although this country has confidently crossed the epidemic threshold. Also, it is not yet possible to analyse the situation in Brazil, since the epidemic in this country is at the initial stage of the exponential growth of the epidemic, despite the unprecedentedly large number of officially recorded cases in this country.

## Conclusion

We have found a regular component of the monitoring error of officially registered total cases of the spread of the current pandemic. This regular error component explains the reason for the failure of a priori mathematical modelling of probable epidemic events in different countries of the world. Processing statistical data of countries that have reached an epidemic peak has shown that this regular monitoring obeys a simple analytical regularity. This pattern allows us to answer the question: is this or that country that has already crossed the threshold of the epidemic close to a peak or is still far from it.

Not far off is that happy day when the world will cope with the pandemic as a whole. Monitoring data on the current pandemic, collected in its entirety, will allow us to establish valuable a posteriori patterns to significantly improve the quality of a priori dynamic modelling of epidemic events when they appear in the future.

The countries of the “golden billion”, despite the sometimes catastrophic situation allowed in this part of the world, judging by the dynamics of current events, are the first to pass the peak of the epidemic. The fate of the rest, most of the world remains uncertain. Special attention of an enlightened society should be focused today on the most painful epidemic situation in the USA, Brazil and Russia. The statistics of these countries indicate the absence of a precedent in the history of the epidemic compared to other countries that are already close to recovery. A country called the World is at the beginning of the pandemic despite the truly gigantic scale of the disaster already achieved.

## Data Availability

https://kovriguineda.ucoz.ru/COVID-19/COVID-19-04.mw

## Footnotes

† The United Kingdom has reached the epidemic peak while writing this text.

